# Estimating health facility-level catchment populations using routine surveillance data and a Bayesian gravity model

**DOI:** 10.1101/2025.02.13.25322240

**Authors:** Justin Millar, Rohan Arambepola, Ewan Cameron, Busiku Hamainza, Kafula Silumbe, John Miller, Adam Bennett, Hannah Slater

## Abstract

Accurate estimates of health facility catchment populations are crucial for understanding spatial heterogeneity in disease incidence, targeting healthcare interventions, and allocating resources effectively. Despite improvements in health facility reporting, reliable catchment population data remain sparse. This study introduces a Bayesian gravity model-based approach for estimating catchment populations at health facilities, with a focus on Zambia’s routine malaria surveillance data from 2018-2023. Our method integrates health-seeking behavior, facility attractiveness, and travel time, allowing for the development of probabilistic catchment areas that reflect the treat-seeking and facility selection process. We developed an open-source R package to implement this method, and we apply this model to Zambian health facilities and compare the results to reported headcount data, highlighting improvements in stratification of malaria incidence rates. Additionally, we validate the model’s sensitivity using real-world treatment-seeking data from household surveys in Southern Province, Zambia, demonstrating its utility in enhancing sub-district-level health facility data for strategic planning. Validation of model facility selection rates compared to the treatment-seeking data showed a model sensitivity of 0.72 overall, with sensitivity reaching 0.89 for households within 2 kilometers of their preferred facility. This validation supports the model’s ability to closely estimate treatment-seeking behavior patterns, offering a scalable, accurate tool for enhancing local-level decision-making for health interventions, contributing to improved targeting and understanding of healthcare access patterns.

## Introduction

As routine disease surveillance systems improve worldwide, there is an increasing demand to use these data to understand local-level heterogeneity in disease incidence so that interventions can be targeted to the most at-risk populations. Calculating disease incidence rates at the health facility level requires accurate estimates of both the number of confirmed cases and the number of people in the facility’s catchment population. In recent years, improved reporting from health facilities to national health management information systems (HMISs), wider availability of diagnostic tools (such as rapid diagnostic tests for malaria), and increased healthcare access have improved the representativeness of these data (1–3). In contrast, accurate catchment population sizes are often unavailable at the health facility level, even for well-developed HMISs (4–6). There are many aspects to the “denominator challenge”, including a lack of standardized processes for defining, estimating, and documenting catchment populations for individual health facilities, which can be maintained and updated over time (7–9). As a result, it can be difficult or impossible to calculate accurate incidence rates that could be used to understand heterogeneity in disease risk, intervention targeting, and commodity needs at the health facility catchment level.

Therefore, national programs require standardized methods and tools for estimating health facility catchment population in order to support local-scale decision making. These estimates should incorporate characteristics of health-seeking behaviors, such as individual facility characteristics/competition, the time it takes to travel to a facility based on a given mode of transport, and individual preference to, for example, bypass the nearest facility (10). Additionally, methods to estimate catchment populations should be well-documented and straightforward to update over time to account for population growth or facilities opening or closing.

Several approaches for estimating health facility catchment populations are available. Ad hoc methods, such as distributing a district population total among facilities in proportion to the number of malaria cases or outpatient visits at each facility, may obscure local-level heterogeneity. Approaches that rely on the local community to draw geographic catchment boundaries on settlement maps have been used in Zambia and elsewhere(e.g., (11,12)), however, they are time and resource intensive and may not be scalable or easy to keep updated. They also may not account for the inherent human behavior aspects of where individuals choose to seek care.

Several quantitative modelling approaches have been used to estimate catchment population, with varying degrees of data requirement and complexity tradeoffs. Spatial buffering, drawing a fixed radius around a facility location and allocating all people within the radius to the facility’s catchment, is a commonly used method due to its ease of use and implementation (13,14). However it has several limitations including requiring an (often arbitrary) radius choice, not accounting for travel time, the potential for overlapping buffer areas, which may lead to double counting of populations, and not accounting for differences between individual health facilities besides location. Areal delineation techniques, such as Voronoi tessellation, are easy to implement in GIS software and can prove useful for planning and implementing intervention campaigns, however, they imply that all individuals in a given area seek always treatment from the same facility and can also produce unrealistically large or small catchment areas in rural or urban areas, respectively. More complex methods, such as network flow analysis (15,16), two-step floating catchment (17–20), and the Huff model of spatial attractiveness (21–23), can provide a more accurate representation of treatment-seeking, human movement, and resource utilization rates. However, these methods are dependent on specific, high-quality data, such as transportation networks, geocoded patient data, supply or quality of healthcare providers, and socio-economic indicators, which may not be readily available and thus unsuitable for low-resource settings (24).

Statistically derived treatment seeking probability surfaces, such as gravity models, have gained attention as a tool for estimating facility catchment populations (25,26). These models combine health facility data with population and travel time information to derive treatment-seeking estimates for all locations, which can then be used to calculate population denominators for individual facilities.

In this study, we implement a gravity model-based statistical method for estimating health facility catchment populations. This approach considers the spatial distribution of the population, travel times to health facilities, and facility attractiveness (27,28). This method implicitly allows for overlapping catchment areas, meaning that individuals residing in a given pixel are assigned probabilities that they will attend each of several nearby health facilities – this results in more realistic estimates of treatment seeking behavior (29–31). We also developed an open-source R package for this method and demonstrate its applications using health facility locations, population data, and confirmed malaria case data from the Zambia HMIS (2018–2023). We compare our catchment population estimates to reported head count data and show how this method can improve sub-district stratification maps based on surveillance data. Additionally, we evaluate model sensitivity using field survey data on household treatment-seeking choices. Finally, we discuss potential use cases and extensions of this method to incorporate more facility-specific information.

## Methods

### Statistical framework

The purpose of a catchment model is to estimate the likelihood that a treatment-seeking individual in a given location would select a particular health facility from a set of possible health facility locations. A facility-specific catchment population can then be calculated by taking the sum of the expected number of individuals from each location over all possible locations.

Our specification of the catchment model framework incorporates two key components of treatment-seeking behavior – estimated travel time and individual facility attractiveness. This model specification is referred to as a “gravity” model, where distance and mass components are represented by the travel time and facility-specific weight which represents its relative attractiveness, respectively.

Geographic distance and travel times are often critical factors in treatment-seeking behavior (32–34). In our catchment model, the initial treatment-seeking probability is based on estimated walking travel time to each facility, such that the likelihood of an individual in location *i* seeking treatment would do so from health facility *j*, *p*(*i* → *j*) is proportional to the inverse of the squared travel time to that facility (similar to (35)). That is,

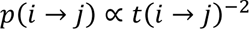

where *t*(*i* → *j*) is the travel time from location *i* to location *j* in minutes. The initial catchment population of health facility *j*, *pop*_HF(*j*)_ is then given by:

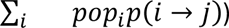

where *pop*_*i*_ is the population in location *i*.

Individual characteristics of health facilities also play an important role in facility selection. Multiple factors, such as facility size and type, public versus private ownership, available services, costs to patients, insurance acceptance, wait times, and professionalism of staff can influence treatment-seeking behavior and may lead an individual to opt for a longer travel time to attend their preferred facility (36–38). Incorporating an exhaustive list of intrinsic facility-level factors is difficult to do on a systematic level (39). In some cases, multiple factors are decomposed into individual facility scores, which are then used to either categorize facilities or in a hierarchical framework (40,41).

An alternative approach is to infer an overall “attractiveness” weight for each health facility, representing the relative effect of facility-specific factors on the probability of individuals seeking treatment at each facility. The facility attractiveness weights are initialized using routine surveillance data, such as the average number of outpatients or malaria cases per month, and then updated by the gravity model. For instance, a health facility with more patients or cases than would be expected based on only travel time would receive a larger attractiveness weight.

Specifically, for any given values of health facility weights, *w*_1_,…, *w*_*n*_, the probability of an individual in location *i* visiting facility *j* was proportional to

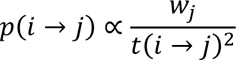

That is, as before this probability depends on the travel time for location *i* to visit facility *j* but this probability is now scaled by the attractiveness of the facility. As before, the implied catchment facility population is given by taking the sum over all locations where individuals live,

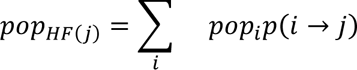

To estimate these weights using the average confirmed malaria cases per month, we model the incidence rate of malaria treatment seeking as a smooth random field over space. Let *r*_*i*_ be this rate in location *i*, then this rate is modelled as

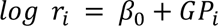

where *β*_0_ is a global scalar parameter to be learned and *GP*_*i*_ is the value of a zero-mean Gaussian process in location *i*. These rates and the probabilities *p*(*i* → *j*) together imply an expected number of malaria cases for health facility *j*,

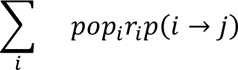

Let *Y*_*j*_ be the average number of malaria cases observed at health facility *j*. Under the assumption that *Y*_*j*_ follows a Poisson distribution, the likelihood for the attractiveness weights, *β*_0_, and the Gaussian process values is given by

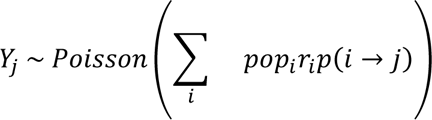

This likelihood is combined with priors on the model parameters to create a full Bayesian model, as well as hyperparameters for learning the flexibility of the Gaussian process and the likely range of attractiveness weights (i.e., how much more or less “attractive” a facility can be relative to other facilities).

We developed an R package called ‘catchment’ to facilitate the implementation of this gravity model framework for estimating health facility catchment populations (42). The catchment package fits the model described above in an approximate Bayesian framework via the INLA and TMB R packages (43,44), which reduces the computational demands compared to the complete Bayesian alternative. The data inputs for the package are a list of health facility locations and initial weights representing attractiveness, a population raster, and a friction surface which is used to calculate distance or estimated travel time between locations. The package is open-source and freely available at https://github.com/PATH-Global-Health/catchment, and provides tools for data processing, model fitting, and extracting and visualizing outputs. More details on implementing the package are provided in the Supplement Materials.

### Source data

Using the catchment R package, we estimated catchment populations for all geolocated health facilities that actively reported malaria cases into the Zambia DHIS2 system from 2018 to 2023. Health facility geolocations were collected from the Zambia DHIS2 system as well as a georepository maintained by the Zambia Ministry of Health (45). Facilities were considered “active” if they reported malaria cases for at least 8 individual months over the study period. Malaria cases passively detected by community health workers were aggregated to their parent facility counts.

The average monthly reported malaria cases were used as the initial value representing facility attractiveness weight *w*_*j*_. We also considered using the average outpatient discharges per month as well as the average non-malaria-related discharges per month, however the outpatient data was lower quality than the malaria data and both metrics were correlated with average malaria cases. As a result, there was little difference in fitted weights between the different initial attractiveness proxy metrics and fewer facilities could be included when incorporating the outpatient data into the attractiveness weight. More details on this comparison are available in the Supplemental Materials.

Population estimates and spatial distribution were based on the 2022 gridded population raster developed by GRID3 (46). This source raster was aggregated to approximately 1 km^2^ resolution. Raster values were then rescaled to approximate the province-level population totals from the 2022 Zambia census (47). We assumed a 3% annual population growth rate to in order estimate populations for the remaining years outside of the census (2018 to 2021, and 2023). This was necessary for deriving facility-level catchment populations for specific years and for the comparison and validation exercises described in subsequent sections.

Walking-based travel time surfaces were created for each health facility using the gridded surface and the non-motorized friction surface developed in Weiss et al. (2020) (48,49). The walking-based friction surface was selected in favor over the motorized friction surface due to the relative low ownership of motorized vehicles in Zambia (3.9% car- and 2.3% motorcycle-ownership nationally) (50). These rasters were resampled to have the same resolution as the population raster, and travel time estimates were based on a least-cost path algorithm using the ‘gdistance’ package in R (51).

### Model fitting

Health facility catchment populations were estimated using the catchment R package following the methodology described above. It was not feasible to fit a single catchment model for the entire country due to the computational constraints and memory demands of large spatial models. Therefore, individual models were fit for each of the 10 provinces. Under this modeling framework individuals could only be allocated to the catchments of health facilities located within their home province, however cross-district travel within a province is possible. To help reduce the memory demands, the models were restricted to only considering the 15 nearest facilities for each pixel based on estimated walking travel time by setting the access probability of further facilities to zero. Extremely remote populations, where the nearest facility was over 6 hours of estimated travel time, were automatically assigned to the nearest facility. Individual INLA meshes were constructed for each model based on province geographies and memory constraints. All other parameters were consistent between each province-level model.

### Comparison and validation

The estimates for health facility catchment populations were compared to the 2022 reported health facility headcounts recorded in the Zambia DHIS2. As described above, the reported headcount data are often incomplete and unreliable, however they are useful for comparing general magnitude and trends with model estimates. Alternative catchment population estimates were derived for each health facility using Euclidean-based Voronoi tessellations and the population raster.

Finally, external validation of model sensitivity was evaluated using household-level data collected in Southern Province, Zambia 2014-15 during a malaria mass test-and-treat (MTAT) campaign (52). These data included household locations, the number of people in each house, and which health facility they reported attending when seeking treatment. Additional details on the validation data and pre-processing steps is described in Supplemental Material. A validation model was fit using the facilities identified in the surveys using the same process illustrated above with the catchment R package. Initial facility-level weights were based on average malaria cases per month per facility reported to DHIS2 from 2014 to 2015. Due to the small geographic area (nine contiguous districts), no additional constraints were required. The model sensitivity was estimated at the pixel-level via multinomial simulation. For each pixel that contained at least one household in the MTAT data, its modeled vector of probabilities was extracted. Based on these probabilities and the population in that pixel from the survey data, a total of 1,000 multinomial draws were taken which resulted in 1,000 predictions of where individuals would seek care. These predictions were then compared to the actual data to calculate the pixel-level sensitivity. This process was repeated across all populated cells and the population-weighted average was calculated to describe the overall model sensitivity.

## Results

### Population estimates and comparisons

Health facility catchment populations were calculated for 2,519 health facilities that reported malaria cases in the Zambia DHIS2 system between 2018 and 2023. First, an access probability surface *p*(*i* → *j*) was produced for each facility using the fitted attractiveness weight *w*_*j*_(Fig. 1). This surface is the estimated likelihood of attending an individual facility from a given pixel. The gravity model only predicts access probabilities for pixels that have non-zero population values, however sparse areas can be filled via imputation to create contiguous polygons (Fig. 1).

**Fig. 1.**
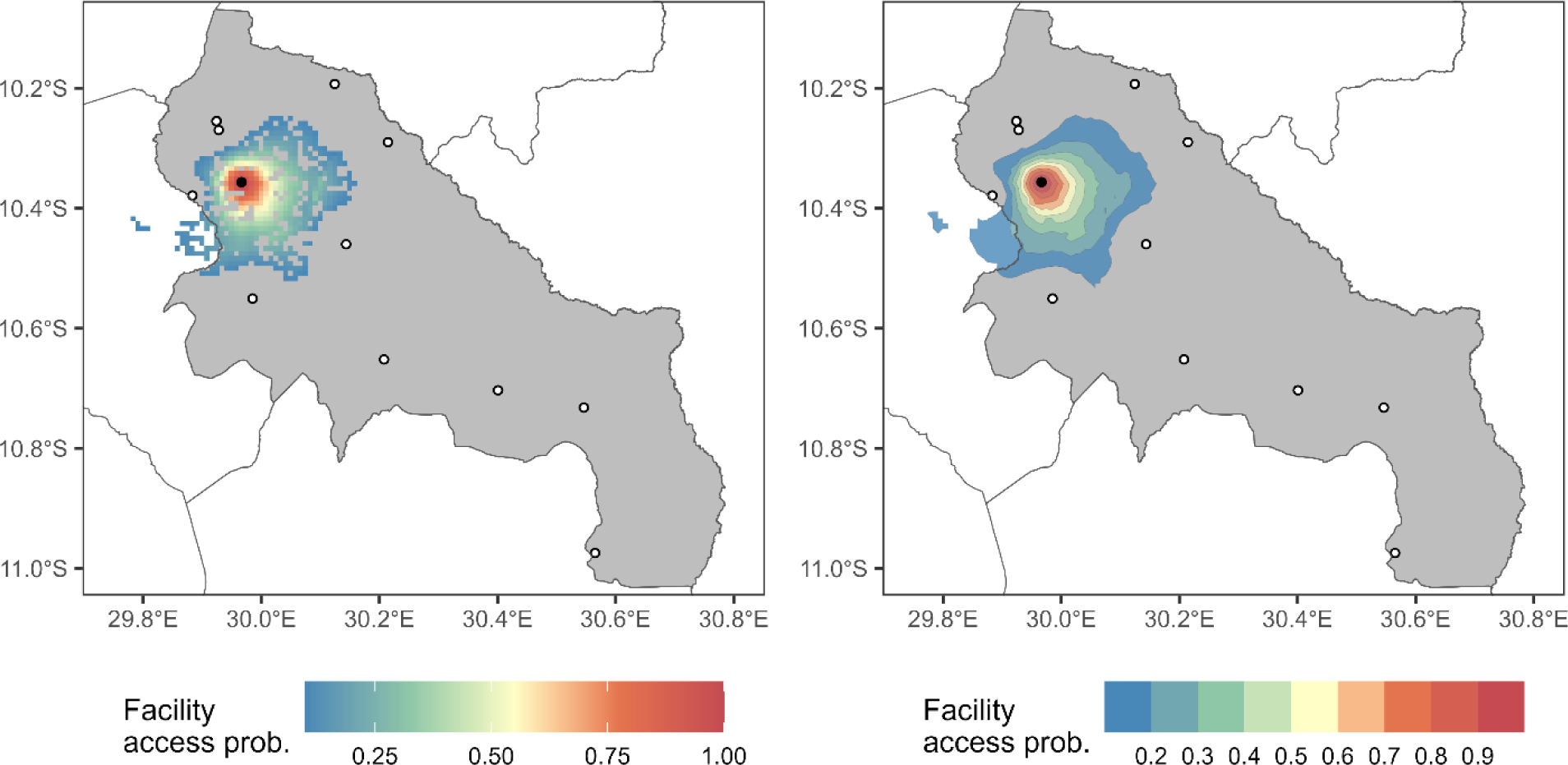
Estimated facility access rate for Nsanja Health Post in Luwingu District. The left panel depicts the estimated likelihood of treatment-seeking individuals attending Nsanja Health Post (black circle), based on their relative distance compared to other neighboring facilities (white circles), and the fitted “attractiveness” weights from the gravity model. Predicted access rates are only generated for populated pixels. The right panel depicts an smoothed interpolated surface where access probabilities have been grouped into binned quantile isobands.

Visualizing the probabilistic catchment areas can help illustrate the similarities and differences with other methods (e.g., Voronoi tessellations) and identify areas that might be served by multiple health facilities (Fig. 2). For instance, the gravity model- and Voronoi-based catchment areas can be similar when a facility is far from the nearest neighboring facilities but may be very different when facilities are closer to each other due to the geographic flexibility of the catchment model (Fig 2). Therefore, differences in population estimates from the Voronoi and gravity model methodologies are likely to be less dramatic in rural areas where health facility locations are spread out, and more extreme in urban and peri-urban areas where there is a higher geographic density of people and health facility locations.

**Fig. 2.**
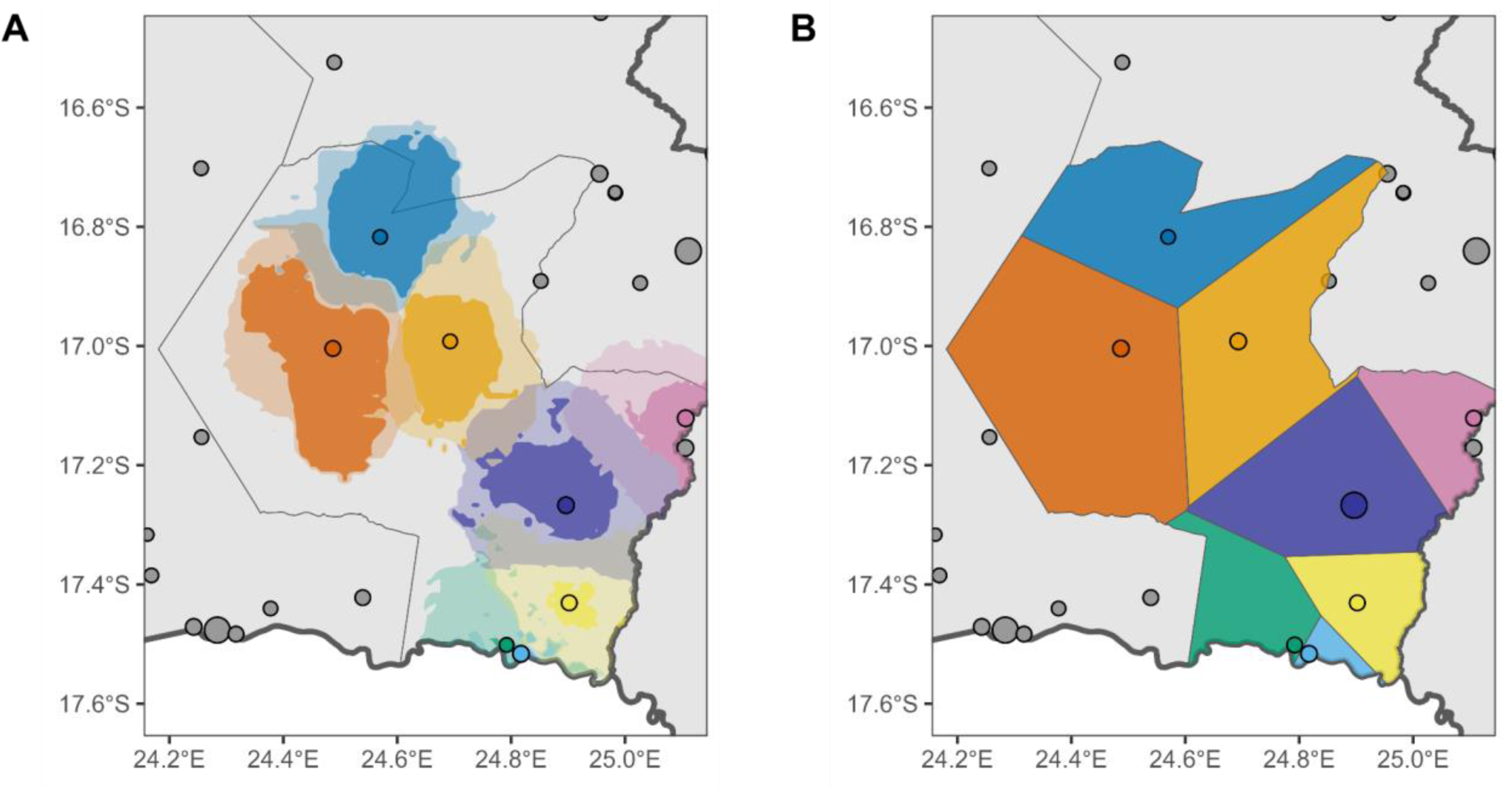
Comparison of health facility catchment areas in Mwandi District. Both panels depicted health facilities in Mwandi district (colored circles), and in neighboring districts (gray circles), and the size of the circle is based on their estimated catchment population. District and province boundaries are depicted by thin and thick lines, respectively. Panel A shows the implied probabilistic catchment area for each facility based on interpolated isobands from the access probability surfaces for each facility in Mwandi. The darker colored polygons contain areas that have a 75% or higher predicted access probability, while the lighter colored areas that have a 25% or higher predicted access probability.

Facility-level catchment populations were generated for each year from 2018 to 2023 based on the facilities that actively reported cases during each year. Each yearly set of facility-level catchment population estimates was based on the adjusted population surface for that year (using the assumed 3% population growth rate) and which health facilities actively reported cases during that specific year.

Most facilities were identified as either health posts (n = 1,031), rural health centers (n = 1,085), or urban health centers (n = 264). The distribution of estimated facility catchment populations by facility type was as follows (Fig. 3); the facility-level populations for health post (median: 4,543, IQR: 2,816 – 6,886) were on average smaller than rural health centers (median: 6,068, IQR: 4,121 – 8,461), which were on average smaller than urban health centers (median: 11,993, IQR: 6,812 – 19,460). Public hospital and hospital-associated health centers (HAHC) (n = 96) comprise large district and regional hospitals, as well as smaller referral hospitals. As a result, this category comprised a wide range of catchment sizes (median: 10,232, IQR: 6,559 – 15,004). Typically patients in Zambia will initially seek care from a close health post or health center before seeking treatment at a hospital. However, hospitals are generally located in higher population-density regions which may cause them to have larger catchment population estimates. The remaining facilities (n = 43) were categorized as “Other”, which comprised of private facilities and facilities that could not be categorized.

**Fig. 3.**
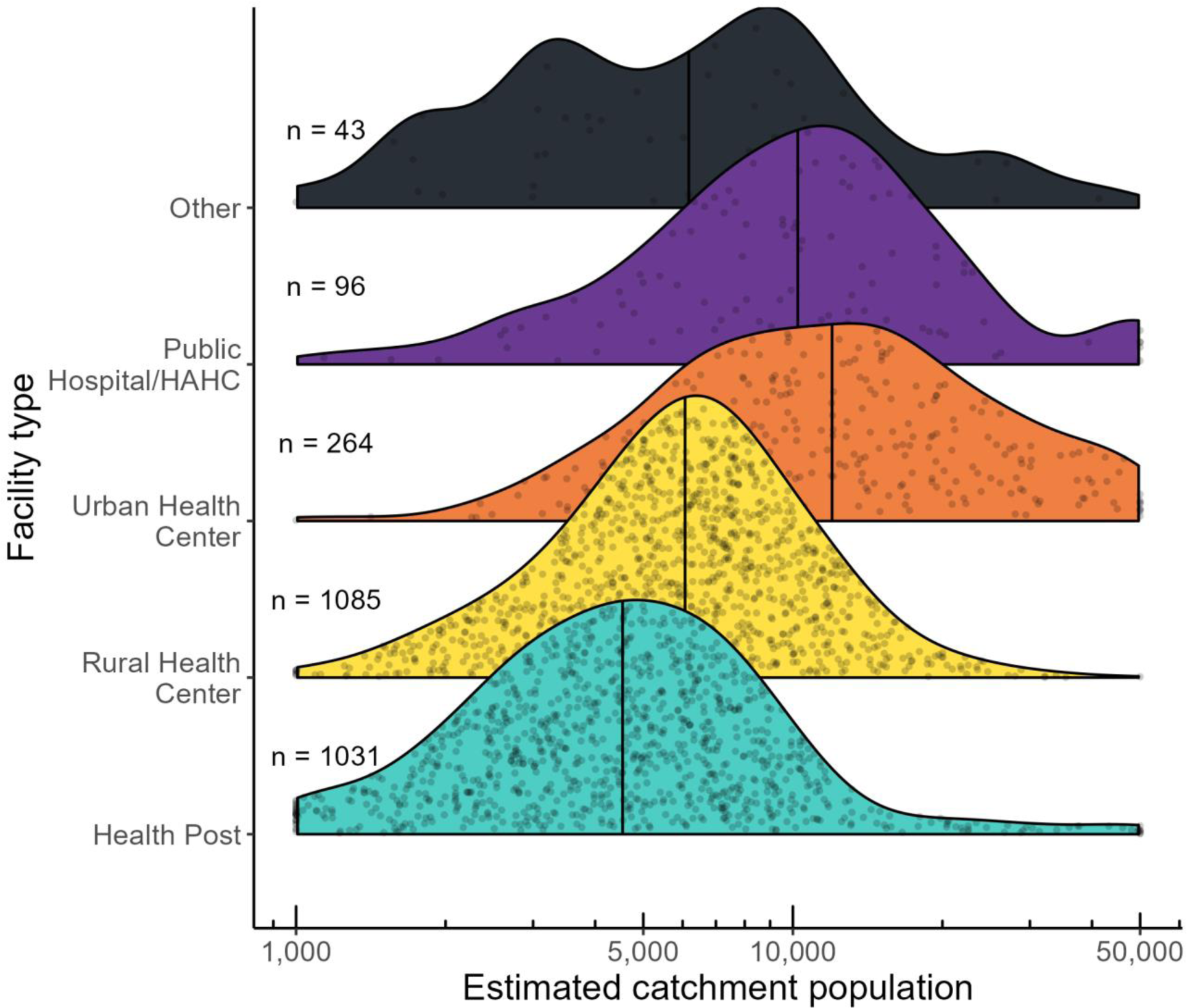
Estimated catchment population size in 2022 by facility type. The center line depicts the median value for each facility type. Individual facilities are represented by points within their respective density curve.

The modeled population estimates were compared to the headcount values that were reported into the Zambia DHIS2 system (Fig. 4). Headcount values based on province-level manager review are often generated using methods that are not well-documented, they are typically not updated regularly, and are often completely missing. Additionally, the sum of facility headcounts is often lower than census areal estimates at the district- and province-level. However, the facility population estimates should follow the same general trends observed in the reported headcount.

**Fig 4.**
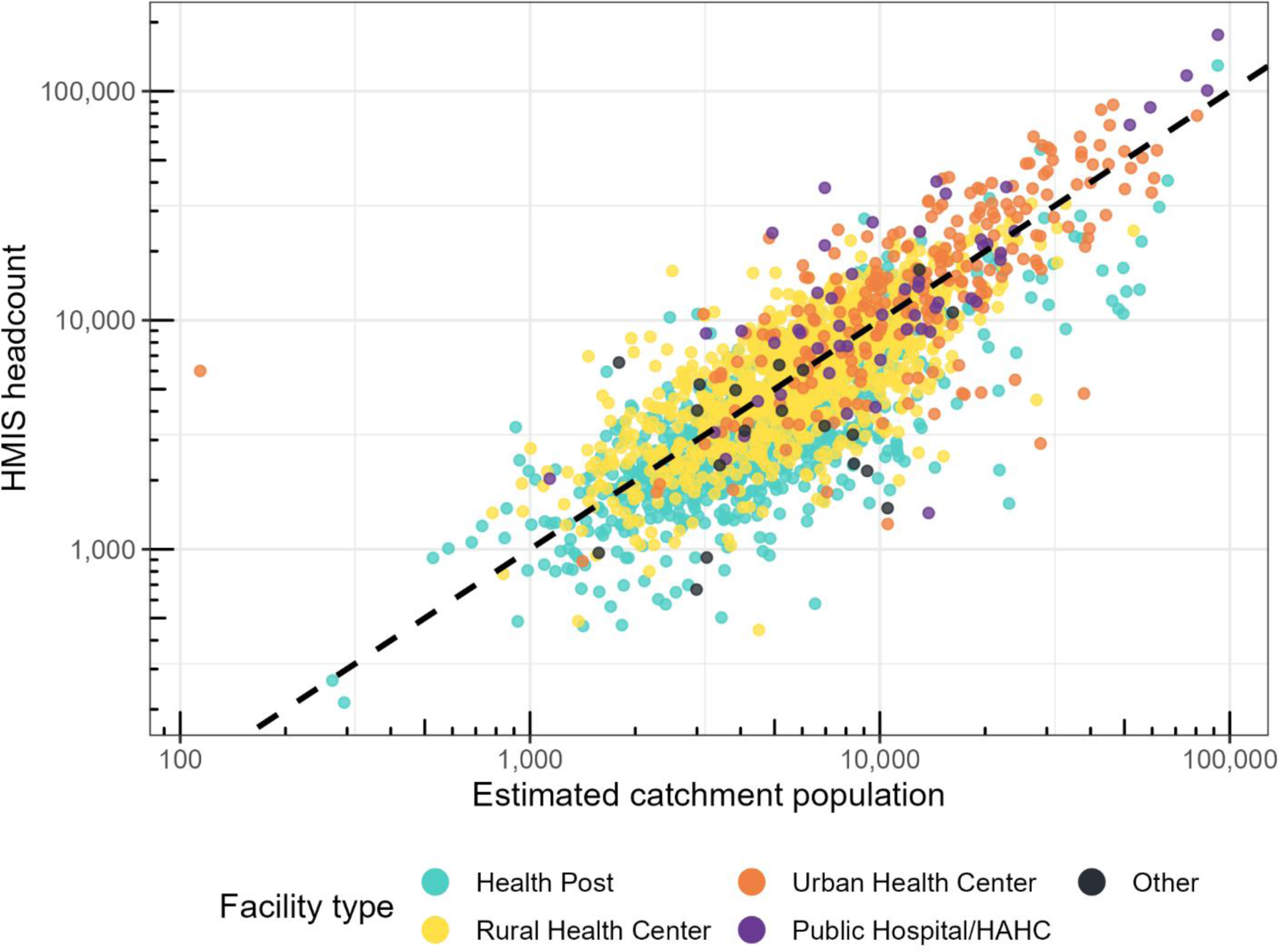
Comparison of estimated facility catchment population and reported headcount. Each point represents a health facility that reported malaria in 2022. The dashed line is a standard x = y line. Total values between the catchment estimates and headcounts differ. This plot does not include facilities where the headcount is missing or reported as zero despite reporting malaria cases.

In 2022, there were 239 facilities that actively reported malaria cases into DHIS2 but did not have recorded headcount data. Among the remaining facilities, there was a high correlation between the reported headcount and the modeled population estimates (R = 0.810). The total reported headcount population for 2022 was 17,333,601, which is 11.6% lower than the 2022 census estimate of 19,610,769. This disparity is more pronounced at the provincial level, where the difference between headcount totals and estimate totals was as high as 22.7% (Table 1). In contrast, the population raster used in the catchment model was scaled to the census values. As a result, the sum of the catchment populations from the modeled estimates is within ±0.2% of the census estimates for each province.

**Table 1.** Comparison of 2022 province population totals.

| Province: | Census<br>population: | Sum of estimated<br>catchment<br>populations from<br>model: | Sum of reported<br>headcount: | Difference between<br>reported<br>headcounts and<br>census: |
| --- | --- | --- | --- | --- |
| Central | 2,252,483 | 2,252,557 | 1,960,766 | -13.0% |
| Copperbelt | 2,757,539 | 2,756,687 | 2,562,322 | -7.1% |
| Eastern | 2,454,788 | 2,452,701 | 2,211,827 | -9.9% |
| Luapula | 1,514,011 | 1,511,133 | 1,280,029 | -15.5% |
| Lusaka | 3,079,964 | 3,078,381 | 2,784,266 | -9.6% |
| Muchinga | 918,296 | 916,291 | 719,302 | -21.7% |
| Northern | 1,618,412 | 1,618,589 | 1,250,837 | -22.7% |
| Northwestern | 1,270,028 | 1,269,672 | 1,067,155 | -16.0% |
| Southern | 2,381,728 | 2,381,719 | 2,205,885 | -7.4% |
| Western | 1,363,520 | 1,361,261 | 1,291,212 | -5.3% |
| TOTAL | 19,610,769 | 19,598,991 | 17,333,601 | -11.6% |

### Estimating malaria incidence rates

Malaria incidence rates were calculated for each health facility from 2018 to 2023 using the total reported cases in DHIS2 and the estimated catchment populations (Fig. 5). The Zambia National Malaria Elimination Program uses reported incidence to categorize health facility catchments into stratification zones which are used for strategic planning and subnational intervention targeting. Annual health facility incidence rates were calculated for each individual year using the year-specific adjusted population raster and the subset of facilities that reported malaria in a given year (Fig. 5).

**Fig. 5.**
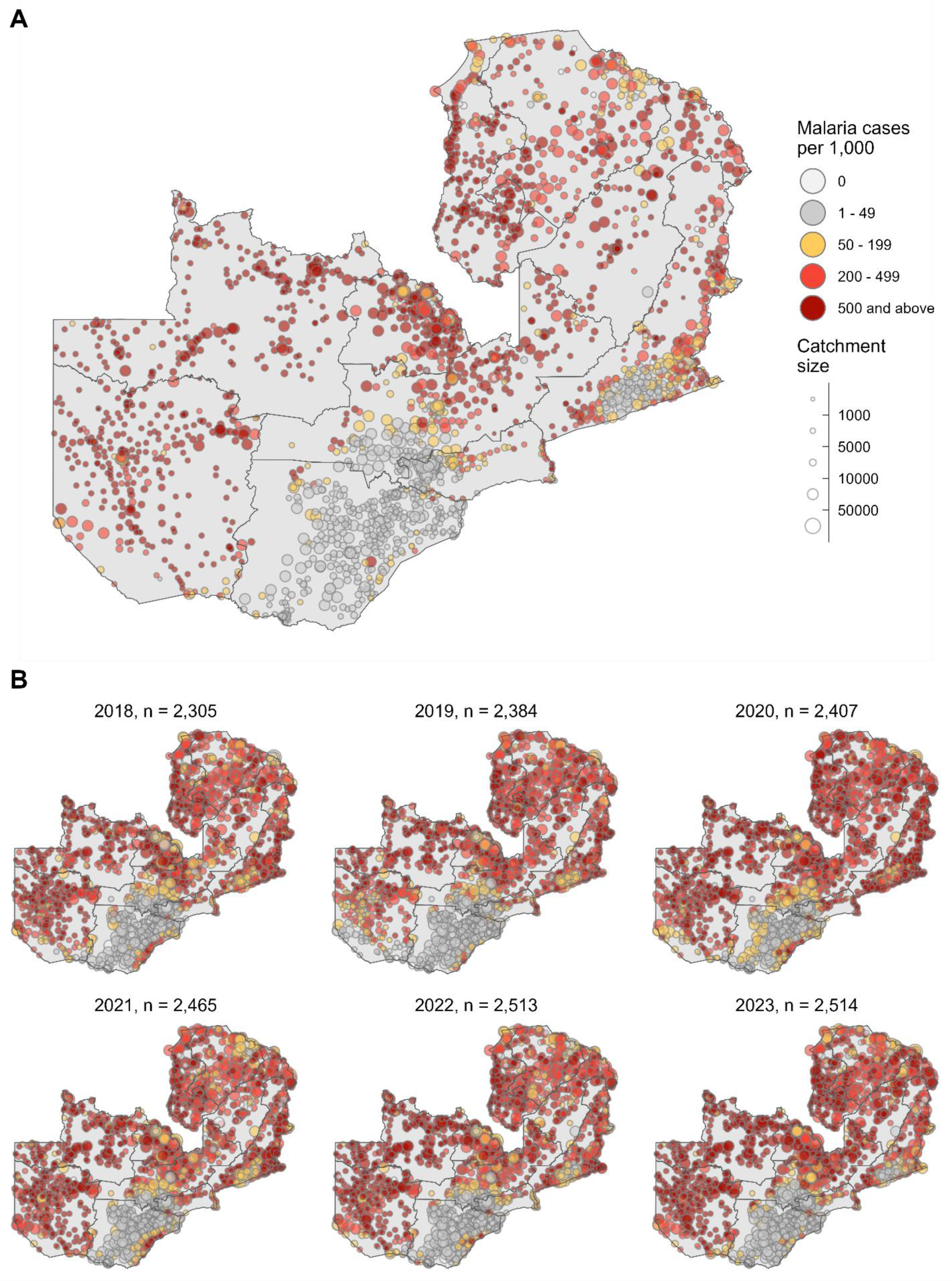
Malaria incidence stratification by health facility catchment area. Panel A depicts incidence rates based on reported malaria cases in 2022 and the estimated catchment population using the gravity model. Panel B depicts the estimated annual incidence rates for all health facilities that reported malaria cases for each year from 2018 to 2023. Incidence categories are based on the defined stratification zones used by the Zambia National Malaria Elimination Program.

Historically, reported headcounts have been used as the population denominator for calculating facility-level malaria incidence rates, however limitations due to missing records and underestimation of the total population can substantially impact the incidence estimates. As a result, the headcount-based incidence stratification differs substantially from the incidence calculated using model-based population estimates. In 2022, there were 732 health facility catchments (29.1% of total reporting facilities) that were placed in different stratification zones when using headcount-based incidence rates compared to the model-based incidence estimates (Fig. 6). In most cases the headcount-based incidence was in a higher stratification zone (n = 575, 78.6%) than the model-based incidence. Additionally, 239 facilities were missing headcount data but could now be stratified using the model-based population estimates.

**Fig. 6.**
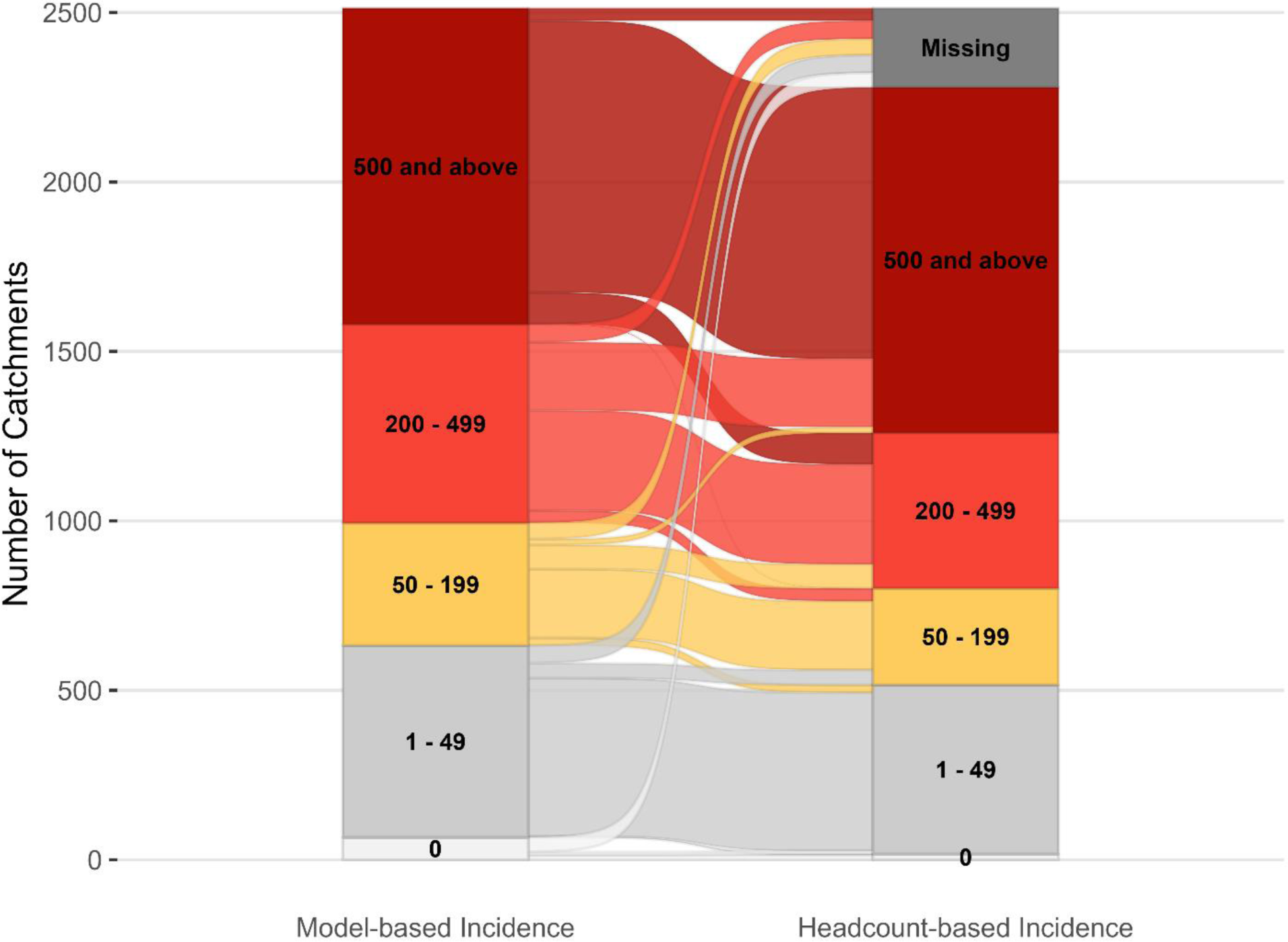
Sankey diagram of the total number of health facility catchments in each malaria incidence zone in Zambia during 2022. Malaria incidence (cases per 1,000) was calculated using the total cases reported as the numerator. The left bar used the model estimated facility catchment populations as the denominator, and the right bar used the reported facility headcount in DHIS2.

### Treatment seeking validation

Household data collected during a mass test and treatment campaign in Southern province from 2014 to 2015 were used to validate the gravity model sensitivity via multinomial simulation based on the predicted access probability (52) (Fig. 7). The survey data set contained 43,483 individual households located in 2,162 distinct 1 km^2^ grid cells and a total of 220,603 people. Using this multinomial method described above to compare the modelled catchment population probabilities to the actual data, we obtain a model sensitivity for all 1 km^2^ pixels that contained surveyed households. The mean population-weighted pixel-level model sensitivity was 0.72. Sensitivity tended to be higher in households that were closer to the health facility that they reported attending, as expected by the gravity model framework. For instance, the mean sensitivity for households that were within 2 kilometers to their reported health facility was 0.89, for households that were 5 to 10 kilometers to their reported health facility was 0.65, and households that were 10 to 20 kilometers to their reported health facility was mean sensitivity was 0.43.

**Fig. 7.**
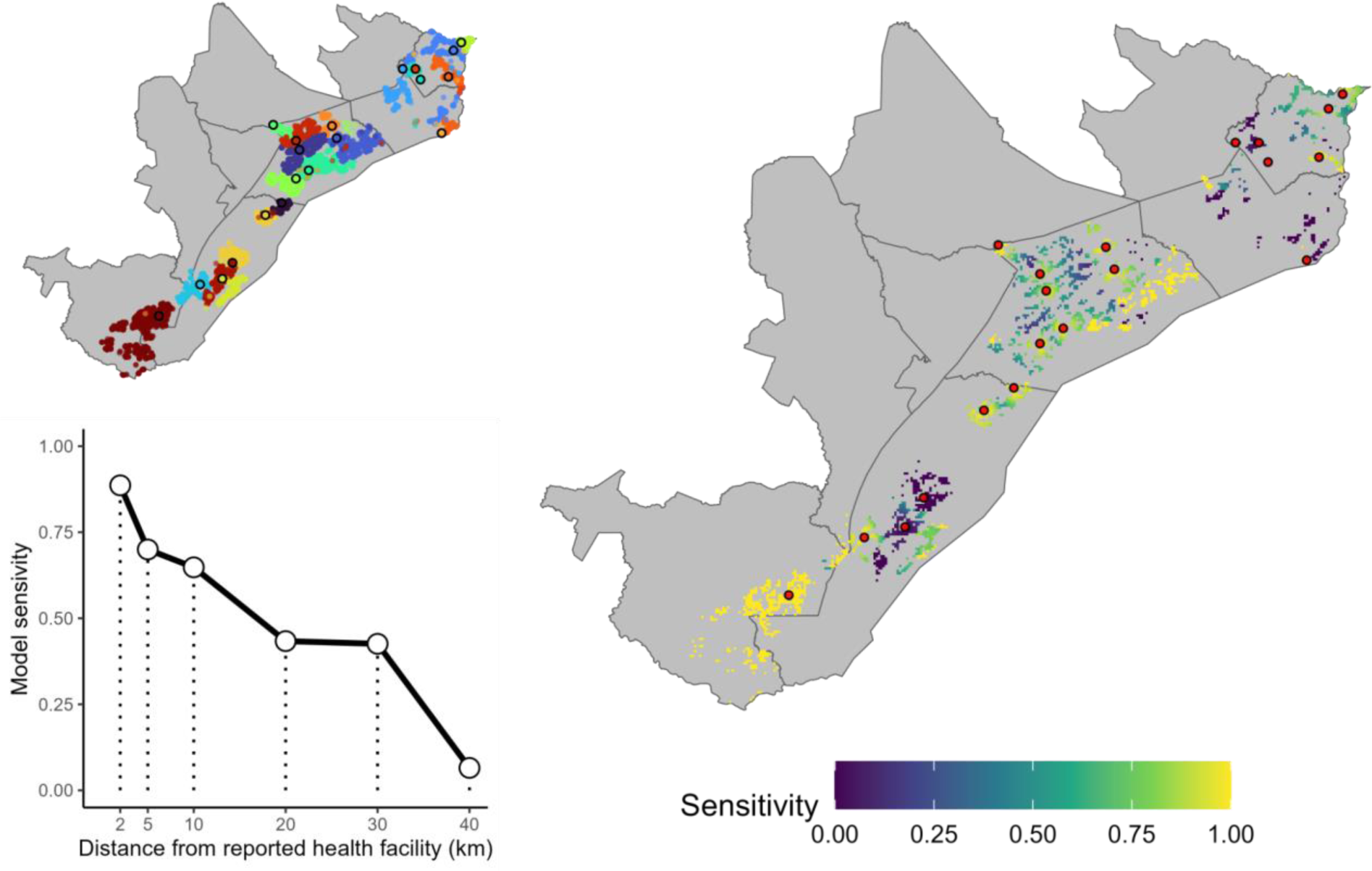
Model validation and sensitivity. The upper left panel shows the locations of households and reported health facility (outlined in black circles) collected in surveys as part of a mass malaria test and treat program conducted in Southern Province, Zambia in 2014-15. Household locations are colored based on which health facility the reported to seek care. The right panel shows the health facility locations (red circles) and pixel-level sensitivity based on multinomial simulation from the predicted facility access probability generated by the gravity-style catchment model. Pixels may contain multiple households. The lower left panel shows the average model sensitivity based on household distance from their reported health facility.

## Discussion

Accurate estimates of health facility catchment populations are essential for understanding local-level variation in disease incidence for intervention targeting, quantifying commodity needs, and guaranteeing that communities have sufficient access to healthcare facilities and healthcare professionals. Current sources of HFCA populations are insufficient – they are often incomplete, the methods used to calculate them are poorly documented, and they are not updated frequently or are inaccurate.

In this study, we present a geostatistical modeling and validation methodology for defining HFCA populations based on a combination of proximity to a health facility and a measure of ‘attractiveness’ of the facility. Our method allows for overlapping catchment area boundaries, ensures catchment populations aggregate to provincial census totals, and is easily implemented on a laptop computer using an open-source R package. We found this method to have high sensitivity when compared to a unique population level dataset on health facility choice in Zambia.

A recent literature review of methods for estimating health facility catchments in sub-Saharan Africa identified persistent gaps in high-quality geocoded data and the need for robust statistical methods for deriving “closer-to-reality” catchment areas and populations (10). Macharia et al. (10) highlighted characteristics that robust methods should contain, including realistic travel time based metrics that incorporate land-cover (rather than Euclidean distance), overlapping catchments, cross-board utilization, and competition between facilities. The review identified studies with similar characteristics as our model (53–55), including those using gravity models (56), however they did not include standardized software packages or code.

The gravity model approach presented in this article incorporates many desired features for a “closer-to-reality” estimate of facility level catchment populations, and our open-source R package allows this framework to be broadly applied. In the absence of complete geocoded patient data, our methodology and software can be used to generate robust “bottom-up” catchment population estimates. Additionally, the population estimates can be dynamically updated to adjust for inactive facilities without needing to refit the model. This can be helpful for estimating population redistribution following service disruption (e.g., long-term stockouts, facility closures, inaccessibility due to flooding or conflict, etc.).

There are however limitations to this approach. Firstly, it requires a complete geo-registry of all health facility locations – if a facility does not have a set of coordinates, it will not be assigned a population in this approach. If there is substantial missingness in the geo-registry, this can lead to overinflated catchment population estimates for facilities that do have coordinates. This method also requires an accurate gridded population raster – there are several publicly available commonly used population rasters (list here…(57–60)), but they all differ slightly which can result in quantitative differences when used to estimate health indicators (61,62). Gridded population surfaces have also been shown to severely underestimate the size of populations living in slums and dense urban areas (63–65)– these can be some of the most at-need populations for health care, so underestimating these populations could result in overburdened and under resourced urban health facilities.

District or province level population estimates extracted from gridded population rasters often do not match the estimates used by MoH – this can lead to discrepancies when incidence rates or commodity needs are summarized at a larger spatial scale. One solution to this is to multiply each pixel in a gridded raster by a district-, province- or national-specific value such that the new aggregated total population matches that used by the MoH. Additionally, in some situations, we may want to produce annual health facility catchment population estimates which would require annual population rasters that account for both population growth and rates of urbanization over time(66).

An important limitation of this modelling approach is that it is difficult to validate the results against real-world data. In this study we have described two validation approaches – one method is to simply compare the modeled population estimates to the HMIS estimated headcounts – and although we suspect the HMIS estimates are inaccurate, observing values of a similar order of magnitude indicates that our estimates are broadly realistic. Additionally, within this validation plot we shade the data points based on the facility type – and we would expect to see catchment population estimates differ based on the facility type – for example health posts should have the lowest catchment populations. Fortuitously, we were able to apply a second validation approach to the results of this model: this involved a large, unique dataset on treatment seeking behavior from several districts in southern Zambia. This allowed us to compare information on which facility individuals sought treatment at compared to their modelled probabilities of attending their nearby facilities. In many situations, such a dataset would not be available and novel validation approaches would be needed This method for estimating catchment population accounts for a key feature of treatment seeking: that individuals may exhibit preferences for different facilities, and these preferences may vary between individuals in a village or even a household (67). In addition this method is easy to implement, will produce district or province level population totals in line with national estimates, and can be easily updated as facilities open and close, or populations grow. Although this example focuses on malaria incidence, the method could be easily applied to other health areas. As health programs increasingly seek to characterize disease trends or evaluate the impact of interventions at the health facility level, these methods will hopefully contribute to more robust and complete estimates to improve the targeting of interventions and understanding changes in risk and disease transmission.

## Data Availability

Model package presented in the article are available at https://github.com/PATH-Global-Health/catchment. Training data for example presented in the article are available upon reasonable request to the authors.

## Acknowledgements

We would like to recognize Pete Gething, Katerine Battle, and Tim Lucas for providing insight and guidance on early version of model and package development. We also want to thank the Zambia National Malaria Elimination Program for supporting this work and use of routine data. We also want to recognize WorldPop, GRID3, and the Malaria Atlas Project for their contributions to open science and providing public resources that were used in this research. This research was funding by the Gates Foundation (grant ID INV-056441).

## Supporting information

The methodology used in this study have been incorporated into an open-source R package which is available at https://github.com/PATH-Global-Health/catchment. This repository contains a tutorial script for reproducing the model described in this article.

## References

1. Bervell B, Al-Samarraie H. A comparative review of mobile health and electronic health utilization in sub-Saharan African countries. Soc Sci Med. 2019 Jul 1;232:1–16.

2. Alegana VA, Okiro EA, Snow RW. Routine data for malaria morbidity estimation in Africa: challenges and prospects. BMC Med. 2020 Jun 3;18(1):121.

3. The malERA Consultative Group on Monitoring E. A Research Agenda for Malaria Eradication: Monitoring, Evaluation, and Surveillance. PLOS Med. 2011 Jan 25;8(1):e1000400.

4. Ibrahim NK. Epidemiologic surveillance for controlling Covid-19 pandemic: types, challenges and implications. J Infect Public Health. 2020 Nov 1;13(11):1630–8.

5. Aborode AT, Hasan MM, Jain S, Okereke M, Adedeji OJ, Karra-Aly A, et al. Impact of poor disease surveillance system on COVID-19 response in africa: Time to rethink and rebuilt. Clin Epidemiol Glob Health. 2021 Oct 1;12:100841.

6. Tatem AJ. Small area population denominators for improved disease surveillance and response. Epidemics. 2022 Dec 1;41:100641.

7. Victora C. What’s the denominator? The Lancet. 1993;342(8863):97–9.

8. Kelley AS, Bollens-Lund E. Identifying the Population with Serious Illness: The “Denominator” Challenge. J Palliat Med. 2018 Mar;21(S2):S-7.

9. Morrison CN, Rundle AG, Branas CC, Chihuri S, Mehranbod C, Li G. The unknown denominator problem in population studies of disease frequency. Spat Spatio-Temporal Epidemiol. 2020 Nov 1;35:100361.

10. Macharia PM, Odhiambo JN, Mumo E, Maina A, Giorgi E, Okiro EA. Approaches to defining health facility catchment areas in sub-Saharan Africa [Internet]. medRxiv; 2022 [cited 2024 Jul 10]. p. 2022.08.18.22278927. Available from: https://www.medrxiv.org/content/10.1101/2022.08.18.22278927v1

11. Kyomuhangi I, Andrada A, Mao Z, Pollard D, Riley C, Bennett A, et al. Assessing national vector control micro-planning in Zambia using the 2021 malaria indicator survey. Malar J. 2023 Nov 30;22(1):365.

12. Stresman GH, Stevenson JC, Owaga C, Marube E, Anyango C, Drakeley C, et al. Validation of three geolocation strategies for health-facility attendees for research and public health surveillance in a rural setting in western Kenya. Epidemiol Infect. 2014 Sep;142(9):1978–89.

13. Lu H, Holt JB, Cheng YJ, Zhang X, Onufrak S, Croft JB. Population-based geographic access to endocrinologists in the United States, 2012. BMC Health Serv Res. 2015 Dec 7;15(1):541.

14. Verma VR, Dash U. Geographical accessibility and spatial coverage modelling of public health care network in rural and remote India. PloS One. 2020;15(10):e0239326.

15. Macharia PM, Ray N, Gitonga CW, Snow RW, Giorgi E. Combining school-catchment area models with geostatistical models for analysing school survey data from low-resource settings: Inferential benefits and limitations. Spat Stat. 2022 Oct 1;51:100679.

16. Huerta Munoz U, Källestål C. Geographical accessibility and spatial coverage modeling of the primary health care network in the Western Province of Rwanda. Int J Health Geogr. 2012 Sep 17;11(1):40.

17. Luo W, Wang F. Measures of Spatial Accessibility to Health Care in a GIS Environment: Synthesis and a Case Study in the Chicago Region. Environ Plan B Plan Des. 2003 Dec 1;30(6):865–84.

18. Delamater PL. Spatial accessibility in suboptimally configured health care systems: A modified two-step floating catchment area (M2SFCA) metric. Health Place. 2013 Nov 1;24:30–43.

19. Shao Y, Luo W. Supply-demand adjusted two-steps floating catchment area (SDA-2SFCA) model for measuring spatial access to health care. Soc Sci Med. 2022 Mar 1;296:114727.

20. Chen X, Jia P. A comparative analysis of accessibility measures by the two-step floating catchment area (2SFCA) method. Int J Geogr Inf Sci. 2019 Sep 2;33(9):1739–58.

21. Luo J. Integrating the Huff Model and Floating Catchment Area Methods to Analyze Spatial Access to Healthcare Services. Trans GIS. 2014;18(3):436–48.

22. Jia P, Wang F, Xierali IM. Using a Huff-Based Model to Delineate Hospital Service Areas. Prof Geogr. 2017 Oct 2;69(4):522–30.

23. Subal J, Paal P, Krisp JM. Quantifying spatial accessibility of general practitioners by applying a modified huff three-step floating catchment area (MH3SFCA) method. Int J Health Geogr. 2021 Feb 17;20(1):9.

24. Macharia PM, Ray N, Giorgi E, Okiro EA, Snow RW. Defining service catchment areas in low-resource settings. BMJ Glob Health [Internet]. 2021 [cited 2024 Apr 9];6(7). Available from: https://www.ncbi.nlm.nih.gov/pmc/articles/PMC8728360/

25. Zinszer K, Charland K, Kigozi R, Dorsey G, Kamya MR, Buckeridge DL. Determining health-care facility catchment areas in Uganda using data on malaria-related visits. Bull World Health Organ. 2014 Mar 1;92(3):178–86.

26. Kapwata T, Manda S. Geographic assessment of access to health care in patients with cardiovascular disease in South Africa. BMC Health Serv Res. 2018 Mar 22;18(1):197.

27. Arambepola R, Keddie SH, Collins EL, Twohig KA, Amratia P, Bertozzi-Villa A, et al. Spatiotemporal mapping of malaria prevalence in Madagascar using routine surveillance and health survey data. Sci Rep. 2020 Oct 22;10(1):18129.

28. Arambepola R, Gething P, Cameron E. Nonparametric Causal Feature Selection for Spatiotemporal Risk Mapping of Malaria Incidence in Madagascar [Internet]. arXiv; 2021 [cited 2024 Apr 9]. Available from: http://arxiv.org/abs/2001.07745

29. Alegana VA, Maina J, Ouma PO, Macharia PM, Wright J, Atkinson PM, et al. National and sub-national variation in patterns of febrile case management in sub-Saharan Africa. Nat Commun. 2018 Nov 26;9(1):4994.

30. Kizito J, Kayendeke M, Nabirye C, Staedke SG, Chandler CI. Improving access to health care for malaria in Africa: a review of literature on what attracts patients. Malar J. 2012 Feb 23;11(1):55.

31. Hjortsberg C. Why do the sick not utilise health care? The case of Zambia. Health Econ. 2003;12(9):755–70.

32. Peters DH, Garg A, Bloom G, Walker DG, Brieger WR, Hafizur Rahman M. Poverty and Access to Health Care in Developing Countries. Ann N Y Acad Sci. 2008 Jun;1136(1):161–71.

33. Noor AM, Amin AA, Gething PW, Atkinson PM, Hay SI, Snow RW. Modelling distances travelled to government health services in Kenya. Trop Med Int Health. 2006 Feb;11(2):188–96.

34. Schoeps A, Gabrysch S, Niamba L, Sié A, Becher H. The Effect of Distance to Health-Care Facilities on Childhood Mortality in Rural Burkina Faso. Am J Epidemiol. 2011 Mar 1;173(5):492–8.

35. Alegana VA, Wright JA, Pentrina U, Noor AM, Snow RW, Atkinson PM. Spatial modelling of healthcare utilisation for treatment of fever in Namibia. Int J Health Geogr. 2012 Feb 15;11(1):6.

36. Tenkorang EY. Health Provider Characteristics and Choice of Health Care Facility among Ghanaian Health Seekers. Health Syst Reform. 2016 Apr 2;2(2):160–70.

37. Escamilla V, Calhoun L, Winston J, Speizer IS. The Role of Distance and Quality on Facility Selection for Maternal and Child Health Services in Urban Kenya. J Urban Health. 2018 Feb 1;95(1):1– 12.

38. Govender K, Girdwood S, Letswalo D, Long L, Meyer-Rath G, Miot J. Primary healthcare seeking behaviour of low-income patients across the public and private health sectors in South Africa. BMC Public Health. 2021 Sep 9;21(1):1649.

39. Field K. Measuring the need for primary health care: an index of relative disadvantage. Appl Geogr. 2000 Oct 1;20(4):305–32.

40. Wang F, Luo W. Assessing spatial and nonspatial factors for healthcare access: towards an integrated approach to defining health professional shortage areas. Health Place. 2005 Jun 1;11(2):131–46.

41. McGrail MR, Humphreys JS. A new index of access to primary care services in rural areas. Aust N Z J Public Health. 2009 Oct 1;33(5):418–23.

42. PATH-Global-Health/catchment [Internet]. PATH; 2024 [cited 2024 Mar 28]. Available from: https://github.com/PATH-Global-Health/catchment

43. Martins TG, Simpson D, Lindgren F, Rue H. Bayesian computing with INLA: New features. Comput Stat Data Anal. 2013 Nov 1;67:68–83.

44. Kristensen K, Nielsen A, Berg CW, Skaug H, Bell BM. TMB: Automatic Differentiation and Laplace Approximation. J Stat Softw. 2016 Apr 4;70:1–21.

45. MFL [Internet]. Ministry of Health Zambia; 2020 [cited 2021 Aug 26]. Available from: https://github.com/MOH-Zambia/MFL

46. GRID3 Zambia Gridded Population Estimates, Version 1.0 [Internet]. [cited 2021 Dec 15]. Available from: https://data.grid3.org/maps/GRID3::grid3-zambia-gridded-population-estimates-version-1-0/about

47. Zambia Statistics Agency. 2022 CENSUS OF POPULATION AND HOUSING PRELIMINARY REPORT [Internet]. 2022. Available from: https://www.zamstats.gov.zm/wp-content/uploads/2023/05/2022-Census-of-Population-and-Housing-Preliminary.pdf

48. Weiss DJ, Nelson A, Vargas-Ruiz CA, Gligorić K, Bavadekar S, Gabrilovich E, et al. Global maps of travel time to healthcare facilities. Nat Med. 2020 Dec;26(12):1835–8.

49. Pfeffer DA, Lucas TCD, May D, Harris J, Rozier J, Twohig KA, et al. malariaAtlas: an R interface to global malariometric data hosted by the Malaria Atlas Project. Malar J. 2018 Oct 5;17(1):352.

50. Zambia National Malaria Indicator Survey 2021 [Internet]. 2021. Available from: https://static1.squarespace.com/static/58d002f017bffcf99fe21889/t/632a39376911de08af7c0ece/1663711548328/Zambia+MIS2021_20220719+SIGNED_for+PRINTING.pdf

51. Etten J van. gdistance: Distances and Routes on Geographical Grids [Internet]. 2018. Available from: https://CRAN.R-project.org/package=gdistance

52. Larsen DA, Bennett A, Silumbe K, Hamainza B, Yukich JO, Keating J, et al. Population-wide malaria testing and treatment with rapid diagnostic tests and artemether-lumefantrine in southern Zambia: a community randomized step-wedge control trial design. Am J Trop Med Hyg. 2015 May;92(5):913–21.

53. Tanser F. Methodology for optimising location of new primary health care facilities in rural communities: a case study in KwaZulu-Natal, South Africa. J Epidemiol Community Health. 2006 Oct 1;60(10):846–50.

54. Stewart K, Li M, Xia Z, Adewole SA, Adeyemo O, Adebamowo C. Modeling spatial access to cervical cancer screening services in Ondo State, Nigeria. Int J Health Geogr. 2020 Jul 21;19(1):28.

55. Gething PW, Noor AM, Zurovac D, Atkinson PM, Hay SI, Nixon MS, et al. Empirical modelling of government health service use by children with fevers in Kenya. Acta Trop. 2004 Aug 1;91(3):227–37.

56. Wilson DP, Blower S. How far will we need to go to reach HIV-infected people in rural South Africa? BMC Med. 2007 Jun 19;5(1):16.

57. Leyk S, Gaughan AE, Adamo SB, de Sherbinin A, Balk D, Freire S, et al. The spatial allocation of population: a review of large-scale gridded population data products and their fitness for use. Earth Syst Sci Data. 2019 Sep 11;11(3):1385–409.

58. Lloyd CT, Sorichetta A, Tatem AJ. High resolution global gridded data for use in population studies. Sci Data. 2017 Jan 31;4(1):170001.

59. High Resolution Population Density Maps + Demographic Estimates by CIESIN and Meta - Registry of Open Data on AWS [Internet]. [cited 2024 Jul 25]. Available from: https://registry.opendata.aws/dataforgood-fb-hrsl/

60. WorldPop. Bottom-up gridded population estimates for Zambia, version 1.0 [Internet]. University of Southampton; 2020 [cited 2024 Jul 25]. Available from: https://data.worldpop.org/repo/wopr/ZMB/population/v1.0/

61. Hierink F, Boo G, Macharia PM, Ouma PO, Timoner P, Levy M, et al. Differences between gridded population data impact measures of geographic access to healthcare in sub-Saharan Africa. Commun Med. 2022 Sep 16;2(1):1–13.

62. Nilsen K, Tejedor-Garavito N, Leasure DR, Utazi CE, Ruktanonchai CW, Wigley AS, et al. A review of geospatial methods for population estimation and their use in constructing reproductive, maternal, newborn, child and adolescent health service indicators. BMC Health Serv Res. 2021 Sep 13;21(1):370.

63. Thomson DR, Gaughan AE, Stevens FR, Yetman G, Elias P, Chen R. Evaluating the Accuracy of Gridded Population Estimates in Slums: A Case Study in Nigeria and Kenya. Urban Sci. 2021 Jun;5(2):48.

64. Thomson DR, Stevens FR, Chen R, Yetman G, Sorichetta A, Gaughan AE. Improving the accuracy of gridded population estimates in cities and slums to monitor SDG 11: Evidence from a simulation study in Namibia. Land Use Policy. 2022 Dec 1;123:106392.

65. Kuffer M, Owusu M, Oliveira L, Sliuzas R, van Rijn F. The Missing Millions in Maps: Exploring Causes of Uncertainties in Global Gridded Population Datasets. ISPRS Int J Geo-Inf. 2022 Jul;11(7):403.

66. article viewer | United Nations iLibrary [Internet]. United Nations; 2018 [cited 2023 Sep 7]. Available from: https://www.un-ilibrary.org/content/books/9789213628829/read

67. Kaboré JMT, Siribié M, Hien D, Soulama I, Barry N, Nombré Y, et al. Attitudes, practices, and determinants of community care-seeking behaviours for fever/malaria episodes in the context of the implementation of multiple first-line therapies for uncomplicated malaria in the health district of Kaya, Burkina Faso. Malar J. 2022 May 30;21(1):155.

